# MuViSS : Muscle, Visceral and Subcutaneous Segmentation by an automatic evaluation method using Deep Learning

**DOI:** 10.1101/2024.03.11.24304074

**Authors:** Edouard Wasielewski, Karim Boudjema, Laurent Sulpice, Thierry Pecot

## Abstract

**Purpose:** Patient body composition is a major factor in patient management. Indeed, assessment of SMI as well as VFA and, to a lesser extent, SFA is a major factor in patient survival, particularly in surgery. However, to date, there is no simple, rapid, open-access assessment method. The aim of this work is to provide a simple, rapid and accurate tool for assessing patients’ body composition.

**Material and methods:** A total of 343 patients underwent liver transplantation at the University Hospital of Rennes between January 1^st^, 2012 and December 31^s^, 2018. Image analysis was performed using the open source software ImageJ. Tissue distinction was based on Hounsfield density. The training dataset used 332 images (320 for training and 12 for validation). The model was evaluated on 11 patients. The complete software and video package is available at https://github.com/tpecot/MuViSS.

**Results:** In total, the model was trained with 332 images and evaluated on 11 images. Model accuracy is 0.974 (SD 0.003), Jaccard’s index is 0.98 for visceral fat, 0.895 for muscle and 0.94 for subcutaneous fat. The Dice index is 0.958 (SD 0.003) for visceral fat, 0.944 (SD: 0.012) for muscle and 0.970 (SD: 0.013) for subcutaneous fat. Finally, the Normalized root mean square error is 0.007 for visceral fat, 0.0518 for muscle and 0.0124 for subcutaneous fat.

**Conclusion:** To our knowledge, this is the first freely available model for assessing body composition. The model is fast, simple and accurate, based on Deep Learning.

**Statements and declarations:** All authors declare no conflict of interest

## INTRODUCTION

Sarcopenia is defined by the European Working Group on Sarcopenia in Older People (EWGSOP) as the progressive and generalized reduction of skeletal muscle mass. Well known in the elderly, its incidence is also high in many chronic diseases such as respiratory [1], renal [2] or liver diseases [3]. Sarcopenia, evaluated by the Sketal Muscule Index (SMI) (ratio between the area of the muscles at L3 level and the height squared) is found to be a risk factor for poor prognosis in surgery, in particular with a decrease in the overall survival of patients undergoing hepatic [4], colorectal [5] or pancreatic surgery [6].

Obesity, a true epidemic on a global scale, is defined by the WHO as an abnormal or excessive accumulation of fat, representing a real health risk. Indeed, 4 million people die each year as a result of being overweight or obese. In addition to the risk of developing chronic diseases such as coronary artery disease [7], hypertension [8], dyslipidemia [9] or NASH [10], obesity is recognized in cancerology as a risk factor for many cancers [11], including 13 with a high level of evidence [12]. Moreover, obesity represents a risk factor for mortality in this population [13]. Although global obesity is strongly correlated with abdominal obesity, these 2 parameters are not always linked. In fact, there are patients who are obese, based on BMI calculation, but not viscerally obese, and vice versa. However, this distinction is relatively difficult to assess in clinical practice [14].

The assessment of body composition is complex. Indeed, the gold standard remains the biophotonic X-ray absorptiometry. However, the rarity, the cost and the method (prospective evaluation) do not allow an evaluation in current practice. Computed Tomography (CT), which is faster and widely used for diagnosis and follow-up, is the tool of choice for retrospective assessment of body composition. Several articles in the literature showed that there is a strong correlation between CT data and body composition [15–18]. Therefore, sarcopenia is assessed by the skeletal muscle index (SMI), division of SMA (skeletal muscle mass) by size squared, on the third lumbar vertebra [19,20]. According to the recommendations [21], the assessment of abdominal obesity involves the measurement of the VFA (Visceral Fat Area) on a CT scan.

Although widely used, there are few freely available tools using DeepLearning to assess SMA (skeletal muscle area), VFA (visceral fat area) and SFA (subcutaneous fat area) simply and quickly. In this study, we provide an automated method for assessing SMA, VFA and SFA at the L3 level.

## MATERIAL AND METHODS

### Patient

A total of 331 patients underwent liver transplantation at the University Hospital of Rennes between January 1^st^, 2012 and December 31^s^, 2018. The evaluation of the SMA (Skeletal Muscul Area), SFA (Subcutaneous Fat Area) and VFA (Visceral Fat Area) were assessed from the last CT scan performed in the 90 days before transplantation. The data used to build the MuViss model comes from another study, the results of which have not yet been published. However, the study in question, from which the data are taken, has been approved by the ethics committee of the University Hospital of Rennes.

### Image Analysis

Analysis was performed on a slice through the third lumbar vertebra. The images were processed using ImageJ version 2.3.0/1.53f (open source) (no image analysis software having demonstrated superiority). The distinction between different tissues was based on the Hounsfield Unit (HU). The threshold of -29 to + 150 HU was used for skeletal muscle, which at L3 included: psoas, erector spinae muscles, squared lumbar muscles, transverse abdominal muscles, internal and external oblique muscles, and rectus abdominis muscles. The threshold of -190 to -30 HU was used for fat. Visceral fat included all intraperitoneal and retroperitoneal fat, including perirenal. Subcutaneous fat included all fat located in the muscle wall and the skin plane.

### Image segmentation

#### Code

##### All the used code is open source

U-Net was [22] coded in Python and used the Python libraries numpy [23], tensorflow [24], keras [25], scipy [26] and imgaug [27]. The MuViSS ImageJ macros are available at https://github.com/tpecot/MuViSS.

#### Training dataset

The training dataset consisted of 320 512 × 512 images while the validation dataset consisted of 12 512 × 512 images. For each image in both datasets, a physician expert manually annotated the backbone, visceral fat, muscle and subcutaneous fat with Annotater [28].

#### Training

Images were normalized with a 1-99.8 quantile. A root mean square prop was used to estimate the parameters of the deep neural network by minimizing a weighted cross entropy loss to handle class imbalance for 25 epochs. A data augmentation to increase the training dataset by a factor of 100 was processed before normalization with the imgaug python library [27] and included flipping, pixel dropout, blurring, noise addition and contrast modifications. The trained model in the format that is directly usable with the CSBDeep plugin [29] in ImageJ is available at https://zenodo.org/record/7990044.

#### Post-processing

416 images were then segmented with the trained model. An ImageJ macro [30, 31] was used to correct segmentations. For both backbone and visceral fat, the largest connected components obtained with the MorpholibJ [32] plugin followed by hole filling were defined as the segmented areas. For both muscle and subcutaneous fat, the largest connected components obtained with the MorpholibJ [32] plugin followed by hole filling with a maximum size of 1000 pixels for holes were defined as the segmented areas.

#### Quantification

Pixels with an intensity defined between -29 and 150 were set as muscle while pixels with an intensity between - 190 and -30 were set as fat. The area of muscle pixels within the segmented muscle region as well as the area of fat pixels within the segmented subcutaneous and visceral fat regions were measured to compute the visceral fat area. The MuViSS ImageJ macro defined to process this task is available at https://github.com/tpecot/MuViSS.

## RESULTS

In total, the model was trained on 320 images and evaluated on 11 images. Figure 1 shows examples of the final result after analysis by the model. Table 1 summarizes the types of scanners used in this study both for training, validation and evaluation datasets. The accuracy and evaluation metrics of the segmentation are summarized in Table 2. Note that our model has an accuracy of 97.4%. In order to use our model, video tutorials are available at https://github.com/tpecot/MuViSS.

**Table 1.** Brand and type of scanner.

|  | **Pourcentage (n)** |
| --- | --- |
| **Toshiba** |  |
| Aquilion | 0,19% (1) |
| Aquilion ONE | 0,38% (2) |
| **GE MEDICAL SYSTEMS** |  |
| BrightSpeed | 31,92% (166) |
| BrightSpeed QX/i | 3,27% (17) |
| Discovery CT750 HD | 2,12% (11) |
| LightSpeed16 | 0,58% (3) |
| LightSpeed | 0,38% (2) |
| LightSpeed Pro 32 | 0,58% (3) |
| LightSpeed VCT | 11,92% (62) |
| Optima CT660 | 0,96% (5) |
| Optima CT520 Series | 0,38% (2) |
| Optima CT540 | 0,58% (3) |
| **Philips** |  |
| Brilliance 16 | 1,15% (6) |
| Brilliance 40 | 0,38% (2) |
| Brilliance 64 | 1,73% (9) |
| Ingenuity CT | 0,58% (3) |
| **SIEMENS** |  |
| Mx8000 | 0,96% (5) |
| Definition AS+ | 0,19% (1) |
| Sensation 64 | 0,58% (3) |
| Perspective | 0,19% (1) |
| SOMATOM Definition AS | 1,35% (7) |
| SOMATOM Definition AS+ | 0,96% (5) |
| **Hitachi Medical Corporation** |  |
| SCENARIA | 0,19% (1) |

**Table 2.** Accuracy and evaluation metrics.

|  | **Average** | **Standard deviation** |
| --- | --- | --- |
| **Accuracy** | 0.974 | 0,003 |
| **Jaccard index** |  |  |
| Backbone | 0,919 | 0,016 |
| Visceral fat | 0,980 | 0,006 |
| Muscle | 0,895 | 0,022 |
| Subcutaneous fat | 0,940 | 0,025 |
| **Dice index** |  |  |
| Backbone | 0,958 | 0,009 |
| Visceral fat | 0,990 | 0,003 |
| Muscle | 0,944 | 0,012 |
| Subcutaneous fat | 0,970 | 0,013 |
| **Normalized root mean square error** |  |  |
| Muscle area | 0,0518 |  |
| Visceral fat area | 0,007 |  |
| Subcutaneous fat area | 0,0124 |  |

**Figure 1.**
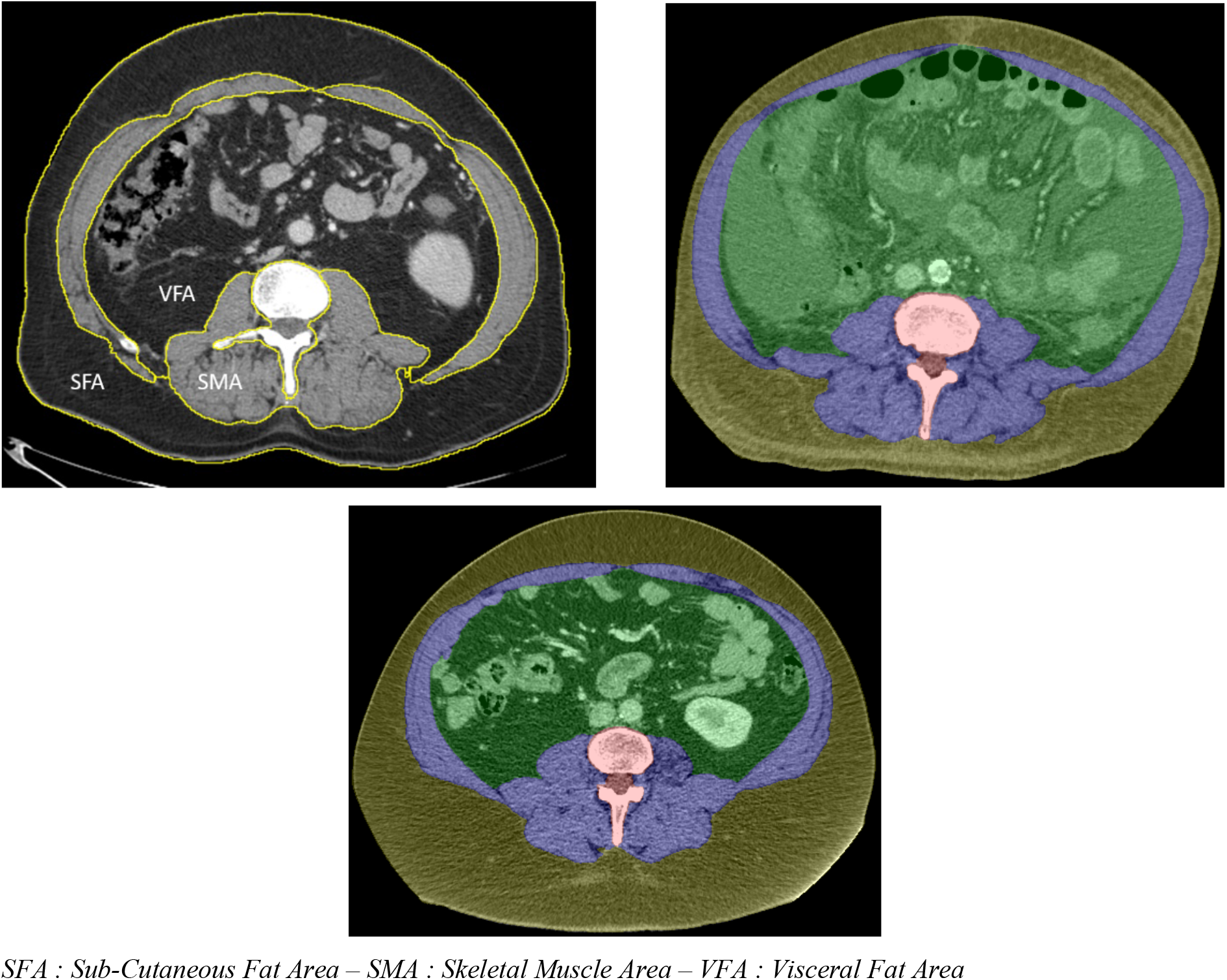
Representation of the automatic segmentation by the model. SFA : Sub-Cutaneous Fat Area – SMA : Skeletal Muscle Area – VFA : Visceral Fat Area

These tutorials show how to install ImageJ and the required plugins and how to use MuViSS. Note that the intensity for muscle, visceral and subcutaneous fat can be adjusted as input parameters.

## DISCUSSION

Sarcopenia and visceral obesity affect a large number of patients, especially in surgery. Their evaluation is often complex and there is currently no simple tool available for their evaluation.

In this study, we demonstrate that a scan-based assessment using Deep Learning was a reliable and reproducible method. This method allows the assessment of the SMA based on the calculation of the area of the L3 level muscles including: the psoas, the erector spinae muscles, the squared lumbar muscles, the transverse abdominal muscles, the internal and external oblique muscles and the rectus abdominis muscles. After adjusting the SMA to the height square, an evaluation of the SMI is made possible. This method also allows to measure, on the same scan section, the VFA represented by the total area of intra-abdominal fat and the SFA represented by the total area of parietal fat.

The evaluation of SMI, a reflection of sarcopenia, is widely studied in the literature. Indeed, in oncology, a 10% loss of SMI in patients undergoing radiochemotherapy for esophageal cancer decreased the overall survival of patients [33]. What’s more, in these patients there is a reduction in progression-free survival and overall survival [34,35]. In the hepatobiliary field, sarcopenia is also a risk factor for 5-year mortality in patients treated for hepatocellular carcinoma. [36]. In gynecological surgery, sarcopenia decreased overall survival in patients with cervical cancer [37], endometrial cancer [38, 39] and ovarian cancer [40]. In pancreatic surgery, there is a decrease in recurrence-free survival and overall survival in patients with low SMI after pancreatic resection [6]. In esophageal surgery, a decrease in survival of sarcopenic patients after esophagectomy has been reported [41]. In colorectal surgery, sarcopenia associated with an NLR < 3 significantly decreased the overall survival of patients operated on for non-metastatic colorectal cancer [42]. In liver transplantation, an increased risk of mortality on the liver transplant waiting list has been highlighted in patients with a low SMI. [43].

The evaluation of obesity is also essential. Indeed, in addition to the specific complications related to obesity such as coronary artery disease, diabetes, etc., obesity, especially visceral obesity, is a risk factor for many complications and a risk factor for mortality. Moreover, although widely used in current practice, BMI is a poor index of fat distribution [44].The assessment of abdominal obesity, represented by the VFA measurement, is much more relevant. Indeed, in pancreatic surgery, visceral obesity associated with sarcopenia leads to a reduction in recurrence-free survival and overall survival. [6]. Moreover, abdominal obesity is a risk factor for postoperative infectious complications after pancreatic surgery [45]. More generally, a meta-analysis summarizes all the postoperative complications associated with abdominal obesity [46].

In the literature, we find a large number of ways of assessing body composition based on scans : BMI_CT [47,48], Aquarius iNtuition (TeraRecon, San Mateo,CA) [49], sliceOmatic (TomoVision, Magog, Quebec, Canada) [50,51], Synapse Vincent [49,50], Infinitt PACS [51]. To date, there has been no significant difference between the different software programs in the assessment of body composition [52]. The aim of this study is not to demonstrate the superiority of our model, which would require a much more in-depth study. However, the advantage of our method is that it’s free. Indeed, although probably very powerful, tools such as SliceOmatic, Aquarius iNtuition, Synapse Vincent and Osirux require a paid license. BMI_CT, on the other hand, is a semi-quantitative evaluation software, meaning that it requires at least one manual segmentation for each evaluation.

The aim was to make available a simple and fast method, free of charge and easy to use. Indeed, our model requires no downloading or special identification, apart from the installation of ImageJ, an open-source software package, or which distributions for Microsoft Windows, Mac OS and Linux are available for download. Our model can therefore be used on any computer. In addition, video tutorials were created to show how to use this tool, available at https://github.com/tpecot/MuViSS.

The interest in developing a free, automatic method could, in future, 1) enable systematic assessment of body composition 2) standardize results and thus enable better comparison.

To date, no team has developed a deep learning-based access-free assessment model for routine body composition assessment. Moreover, the thresholds, in HU, for the evaluation of intra-abdominal and parietal fat is variable according to the studies. Indeed, some teams use the threshold of -190 to -30 HU while others use thresholds of -150 to -50 HU. In this method, the threshold values retained for fat are adjustable, to allow each person to evaluate the VFA according to the chosen values.

## CONCLUSION

To our knowledge, this is the first study to make available a method for evaluating SMA, VFA and SFA using deep learning in free access, based on ImageJ software.

## Data Availability

All data produced in the present work are contained in the manuscript

## Notes

### Competing Interest Statement

The authors have declared no competing interest.

### Funding Statement

This study did not receive any funding

### Author Declarations

Comite d'ethique du CHU de Rennes Approbation du comite d'ethique Avis n 22.161

